# Feasibility of Conducting Comparative Effectiveness Research and Validation of a Clinical Disease Activity Score for Chronic Nonbacterial Osteomyelitis

**DOI:** 10.1101/2022.10.03.22280351

**Authors:** Eveline Y. Wu, Melissa Oliver, Joshua Scheck, Sivia Lapidus, Ummusen Kaya Akca, Shima Yasin, Sara M. Stern, Antonella Insalaco, Manuela Pardeo, Gabriele Simonini, Edoardo Marrani, Xing Wang, Bin Huang, Leonard K. Kovalick, Natalie Rosenwasser, Gabriel Casselman, Adriel Liau, Yurong Shao, Claire Yang, Doaa Mosad Mosa, Lori Tucker, Hermann Girschick, Ronald M. Laxer, Jonathan D. Akikusa, Christian Hedrich, Karen Onel, Fatma Dedeoglu, Marinka Twilt, Polly J. Ferguson, Seza Ozen, Yongdong Zhao

**Author notes:** **Corresponding author:** Yongdong Zhao, MD, PhD, 4800 Sand Point Way NE, Seattle, WA 98105.

## Abstract

**Introduction:** Prospective comparative effectiveness research in chronic nonbacterial osteomyelitis (CNO) is lacking.

**Objectives:** Study objectives were to: 1) determine the use and safety of each consensus treatment plan (CTP) regimen for CNO, 2) the feasibility of using <u>ch</u>ronic nonbacterial <u>o</u>steomyelitis international registry (CHOIR) data for comparative effectiveness research, and 3) develop and validate a CNO clinical disease activity score (CDAS) using CHOIR.

**Methods:** Consenting children or young adults with CNO were enrolled into CHOIR. Demographic, clinical, and imaging data were prospectively collected. The CNO CDAS was developed through a Delphi survey and nominal group technique. External validation surveys were administered to CHOIR participants.

**Results:** 140 (76%) CHOIR participants enrolled between August 2018 and September 2020 received at least one CTP regimen. Baseline characteristics from the three groups were well matched. Patient pain, patient global assessment, and clinical CNO lesion count were key variables included in the CNO CDAS. The CDAS showed a strong correlation with patient/parent report of difficulty using a limb, back, or jaw and patient/parent report of disease severity, but a weak correlation with patient/parent report of fatigue, sadness, and worry. The change in CDAS was significant in patients reporting disease worsening or improvement. The CDAS significantly decreased after initiating second-line treatments from median 12 (8-15.5) to 5 (3-12). While second-line treatments were well tolerated, psoriasis was the most common adverse event.

**Conclusion:** The CNO CDAS was developed and validated for disease monitoring and assessment of treatment effectiveness. CHOIR provided a comprehensive framework for future comparative effectiveness research.

**Key messages:**

- The <u>ch</u>ronic nonbacterial <u>o</u>steomyelitis international registry (CHOIR) provides comprehensive prospective data for comparison of treatment effectiveness
- The clinical disease activity score (CDAS) has content and construct validity to assess CNO

## Introduction

The treatment of chronic nonbacterial osteomyelitis (CNO), also known as chronic recurrent multifocal osteomyelitis (CRMO), has been empirical and based on personal experience, expert opinion, and retrospective observational data (1–10). According to a survey of treating pediatric rheumatologists in 2015, 95% reported the use of nonsteroidal anti-inflammatory drugs (NSAIDs) as first-line therapy for children with CNO (4). While a majority of children with CNO may initially improve with NSAIDs, approximately 50% will have disease worsening within two years (11). Beyond NSAIDs, there was great variability in selecting second-line treatments which most commonly include methotrexate (67%), tumor necrosis factor inhibitors (TNFi) (65%), and bisphosphonates (46%) (4). Large registries of pediatric patients with CNO such as Eurofever (5) and the German National Pediatric Rheumatology Registry (12) have a limited number of CNO patients who received second-line therapies. A retrospective cohort from three tertiary care centers in the United States (8) reported similar efficacy of TNFi compared to corticosteroids in NSAID-refractory patients with CNO, however, bisphosphonates were not evaluated. Recently, a retrospective chart review from Germany and the UK reported that both TNFi and pamidronate were safe and effective in patients with CNO who failed NSAID treatment (13). There appeared to be a quicker treatment response in patients treated with pamidronate, but more sustained remission in the TNFi group (13). Due to lack of robust evidence from prospective international and multi-ethnic studies, supporting the therapeutic efficacy and safety of any of the second-line treatments, the Childhood Arthritis and Rheumatology Research Alliance (CARRA) CNO working group adopted all three categories of treatments as consensus treatment plans (CTPs) for children with CNO who had an NSAID-refractory course and/or with active spinal lesions (14). The standard dosing of treatments and frequency of monitoring were proposed to reduce center-to-center variations for clinical research purpose.

Because of the lack of standardized evaluation tools and sensitive laboratory findings, assessing disease activity in patients with CNO remains a challenge. Historically, pain score has been used as a surrogate for response to therapy, but this approach is limited due to secondary amplified pain syndrome that can occur in CNO (15,16). Magnetic resonance imaging (MRI) can provide detailed information of CNO lesions (17–23) and is used often to evaluate the disease burden for children with CNO at the time of diagnosis. Although regional MRI can be clinically useful, patients can have asymptomatic lesions in other locations. It remains unclear whether all asymptomatic lesions require treatment. Several MRI scoring tools have been developed (18,22,23), but the weighting of lesions at different sites has not been systematically identified. In 2010, Beck et al. (24) developed a core set of CNO outcome variables, PedCNO. Encompassing key variables into a more holistic CNO scale, PedCNO combines patient reported outcomes, laboratory results, and MRI findings. Variables initially thought to be helpful included the erythrocyte sedimentation rate (ESR) and childhood health assessment questionnaire (CHAQ).

In the absence of reliable, prospectively validated and widely accepted disease activity and treatment response criteria, a clinical disease activity assessment tool that is independent of MRI is urgently needed for the assessment of therapies. The <u>ch</u>ronic nonbacterial <u>o</u>steomyelitis international registry (CHOIR) is an international observational longitudinal cohort study of children with CNO that was established by the CARRA CNO workgroup in 2018 (ClinicalTrials.gov Identifier: NCT04725422). The data collection for CHOIR was based on the framework of the CARRA CNO CTPs, and the main target population included children with CNO who failed NSAIDs and/or had high risk lesions (active lesions in vertebral body and/or growth plate) for permanent skeletal damage (14). All the participating sites provided standard clinical care to enrolled patients, and the outcome measurements as well as medication usage were recorded as suggested by the CARRA CNO CTPs (14). The objectives of this study were to: 1) investigate the frequency of use and safety of each CTP regimen, 2) determine the feasibility of using CHOIR data for comparative effectiveness research, and 3) develop and validate a clinical disease activity score (CDAS) for CNO using CHOIR.

## Patients and Methods

### Patient cohort

Children with clinical diagnosis of CNO were enrolled in CHOIR from 14 sites (8 U.S. sites, 2 Canadian sites, 2 Italian sites, 1 Egyptian site, 1 Turkish site) starting August 1, 2018. Informed consent was obtained for all participants and, additionally, assent was obtained from age-appropriate children. Institutional Board Review approval was obtained at each registry site. Inclusion criteria were 1) age at enrollment □21-years-old, 2) presence of bone edema on short tau inverse recovery (STIR) or T2 fat saturation sequence (e.g. TIRM) on MRI within 12 weeks of enrollment, 3) whole body imaging evaluation either by WBMRI or bone scintigraphy, and 4) a bone biopsy to exclude infection or malignancy unless the bone lesions followed a typical CNO distribution or there was concomitant inflammatory bowel disease (IBD), psoriasis, or palmoplantar pustulosis (PPP). Patients were excluded if they had a current malignancy or infectious osteomyelitis and/or any contraindication for conventional synthetic disease-modifying anti-rheumatic drug (csDMARD), TNFi or bisphosphonates. Between the launching date of CHOIR and September 30, 2020, patients who either had NSAID-refractory disease (defined as failure to improve after 1-3 months regular dosing), and/or active spinal lesions (confirmed by MRI), were enrolled and treated according to one of the CTP regimens at the discretion of the treating physician. Patients were not randomized or masked to their treatment assignment. The CTP choices were 1) csDMARD including but not limited to methotrexate and sulfasalazine; 2) TNFi with or without methotrexate; or 3) bisphosphonates including pamidronate or zoledronic acid. Concurrent use of limited courses of NSAIDs and glucocorticoids was permitted for all regimens (14). After October 1, 2021, we expanded enrollment to all children with CNO regardless of their disease status or need for second-line treatments. Our goal was to use patient questionnaires for the external validation of a newly developed CNO CDAS.

### Development of the CNO CDAS

Our workgroup has followed The OMERACT (Outcome Measures in Rheumatoid Arthritis Clinical Trials) filter 2.1 (25) to develop outcome measurements for CNO (26). A scoping review showed life impact and pathophysiological manifestations (26) to be the main core areas reported in the literature. Using this, CDAS domains were discussed at the CARRA CNO work group monthly meetings and narrowed down to physical exam findings, laboratory findings, patient reported outcomes and physician assessment. At the annual CARRA CNO meeting in 2021, a breakout room session and polling as detailed in the results section were conducted to identify and finalize the core variables for the CNO CDAS.

### Validation of the CNO CDAS using CHOIR

Surveys included questions measuring patient reported degrees of difficulty using arms/legs/trunk/jaw, fatigue, depression, worry/anxiety, pain, and patient/parent global assessment using a 0-10 Likert scale. Additional questions (**supplement 1**) assessing self-perceived disease activity (inactive, mildly active, moderately active, or severely active), changes in disease activity compared to the previous visit (improved, worsened, or unchanged), and perceptions of the treatment effectiveness were also administered to participants.

### Data collection

Demographic and disease-related data were collected at baseline visits and follow-up assessments at 3, 6, 9, 12, 18, and 24 months following enrollment. Unscheduled visits occurred if there was a change or adverse event in the CTP medication. Clinical assessments included: 1) patient pain assessment scored on a 0-10 Likert scale; 2) patient/parent global assessment of disease activity scored on a 0-10 Likert scale; 3) physician global assessment of disease activity scored on a 0-10 Likert scale; 4) total number of clinically active CNO lesions defined as focal tenderness, and/or swelling, and/or warmth in addition to patient’s report of pain at a known CNO lesion site (recording sheet is detailed in **supplement 2**); and 5) total CNO lesion count on MRI (14). A CNO CDAS was calculated by summing the patient pain and patient/parent global assessment of disease activity scores and total number of clinically active CNO lesions. Adverse events (AEs) were also collected with special interest on new-onset psoriasis, new-onset autoimmune disease, infection requiring intravenous antibiotics, fracture, and any hospitalization or life-threatening events related to second-line treatments.

### Statistical analyses

Descriptive statistics were used for demographic and clinical characteristics. For continuous variables, medians and interquartile ranges (IQR) were calculated, and for categorical variables, frequencies and percentages were calculated. Mann–Whitney U tests or Fisher’s exact tests were used to assess any significant differences in baseline patient characteristics between CTPs. Log-transformed linear mixed effect models with random participant intercepts were performed to assess CDAS changes after non-NSAID treatments. Wilcoxon signed rank test with continuity correction was performed to determine the change of the CDAS among reported improved, worsened, and unchanged groups. Spearman’s rank correlation test was performed to determine the relationship between the CDAS and reported disease status by patient/families. Participants with missing values or loss to follow-up were excluded from that particular analysis. All computations were performed using R software (version 3.6.1, R Foundation for Statistical Computing, Vienna, Austria). All tests were two-sided, and the results were considered significant if p<0.05.

## Results

### Demographic and clinical data

We identified 140 (78%) out of 179 participants enrolled into CHOIR between August 1, 2018 and September 30, 2020 and treated according to one of the CNO CTP regimens. A total of 39 patients were excluded because of absence of exposure to a second-line treatment. Two received TNFi for pre-existing IBD prior to the diagnosis of CNO. Demographic and clinical features are summarized in **Table 1**. The median age at diagnosis was 9.90 years (IQR 7.99-12.20 years), 55% of the subjects were female, and 84% identified as White Caucasian. There were no significant imbalances in baseline characteristics across three groups including csDMARD-first, TNFi-first (may concomitantly start with methotrexate), and bisphosphonate-first treatment groups (**Table 2**). Specifically, there were no differences between these groups in relation to age, sex, disease duration, interval between diagnosis and treatment start, total MRI lesion count, active spinal lesion, ESR, prior exposure to any csDMARD, and presence of a co-morbid condition including psoriasis and inflammatory bowel disease (IBD). CRP was significantly different among groups (p=0.001).

**Table 1.**
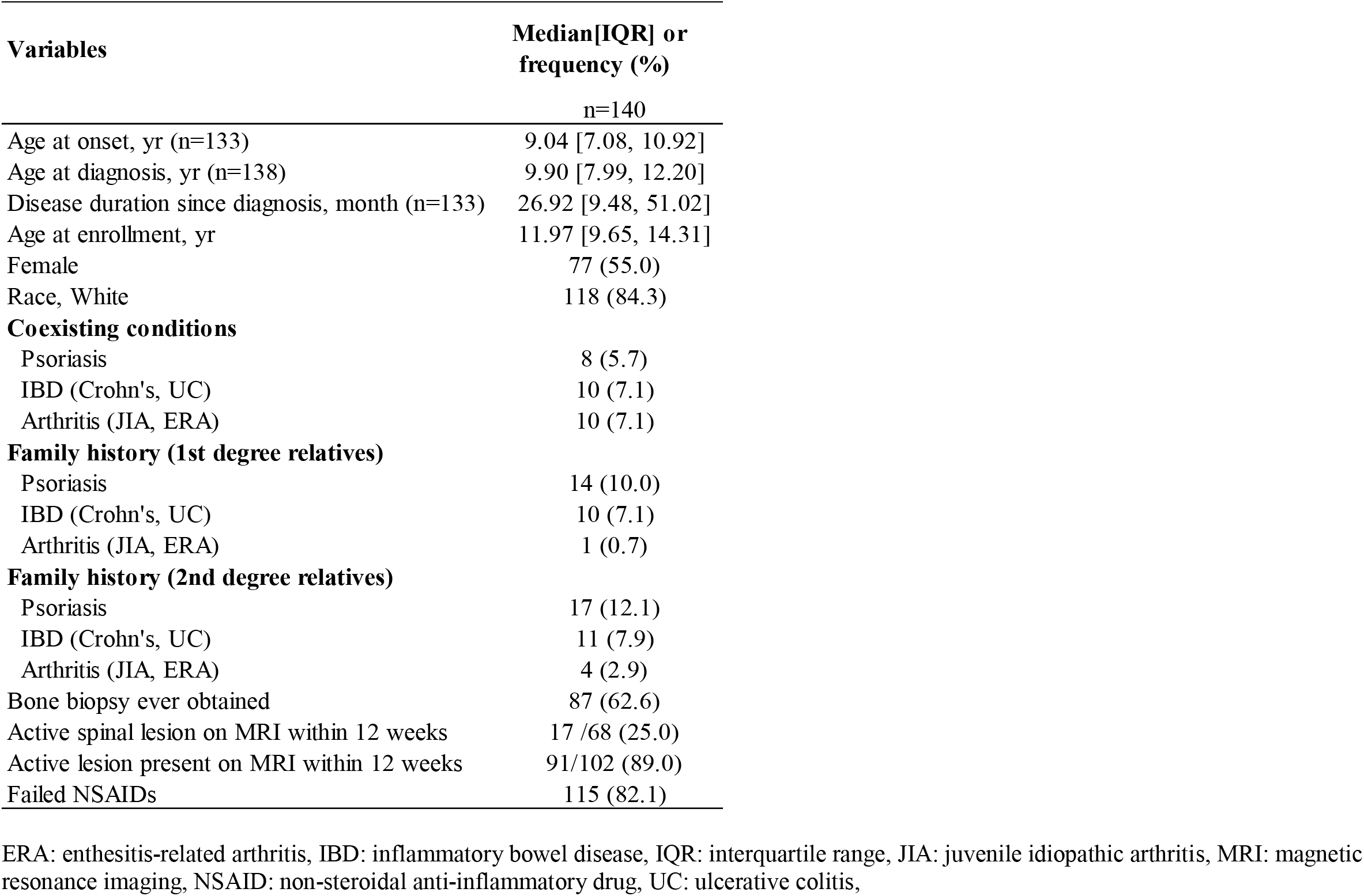
Demographics of the CHOIR inception cohort between 2018 and 2020

| **Variables** | **Median[IQR] or**<br>**frequency (%)** |
| --- | --- |
|  | n=140 |
| Age at onset, yr (n=133) | 9.04 [7.08, 10.92] |
| Age at diagnosis, yr (n=138) | 9.90 [7.99, 12.20] |
| Disease duration since diagnosis, month (n=133) | 26.92 [9.48, 51.02] |
| Age at enrollment, yr | 11.97 [9.65, 14.31] |
| Female | 77 (55.0) |
| Race, White | 118 (84.3) |
| **Coexisting conditions** |  |
| Psoriasis | 8 (5.7) |
| IBD (Crohn's, UC) | 10 (7.1) |
| Arthritis (JIA, ERA) | 10 (7.1) |
| **Family history (1st degree relatives)** |  |
| Psoriasis | 14 (10.0) |
| IBD (Crohn's, UC) | 10 (7.1) |
| Arthritis (JIA, ERA) | 1 (0.7) |
| **Family history (2nd degree relatives)** |  |
| Psoriasis | 17 (12.1) |
| IBD (Crohn's, UC) | 11 (7.9) |
| Arthritis (JIA, ERA) | 4 (2.9) |
| Bone biopsy ever obtained | 87 (62.6) |
| Active spinal lesion on MRI within 12 weeks | 17 /68 (25.0) |
| Active lesion present on MRI within 12 weeks | 91/102 (89.0) |
| Failed NSAIDs | 115 (82.1) |

ERA: enthesitis-related arthritis, IBD: inflammatory bowel disease, IQR: interquartile range, JIA: juvenile idiopathic arthritis, MRI: magnetic resonance imaging, NSAID: non-steroidal anti-inflammatory drug, UC: ulcerative colitis,

**Table 2.**
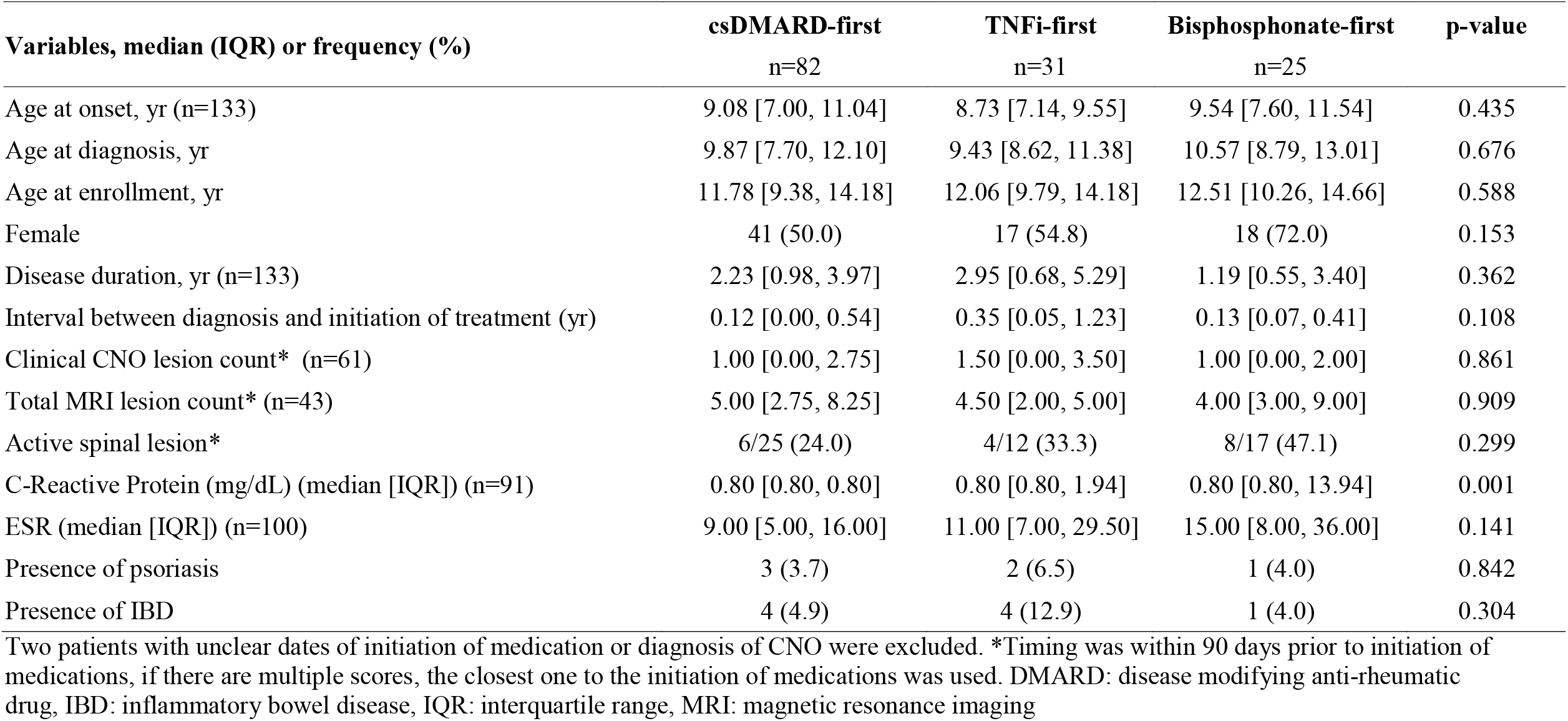
Comparison of pre-treatment characteristics among the three CNO CTP regimens

| **Variables, median (IQR) or frequency (%)** | **csDMARD-first**<br>**n=82** | **TNFi-first**<br>**n=31** | **Bisphosphonate-first**<br>**n=25** | **p-value** |
| --- | --- | --- | --- | --- |
| Age at onset, yr (n=133) | 9.08 [7.00, 11.04] | 8.73 [7.14, 9.55] | 9.54 [7.60, 11.54] | 0.435 |
| Age at diagnosis, yr | 9.87 [7.70, 12.10] | 9.43 [8.62, 11.38] | 10.57 [8.79, 13.01] | 0.676 |
| Age at enrollment, yr | 11.78 [9.38, 14.18] | 12.06 [9.79, 14.18] | 12.51 [10.26, 14.66] | 0.588 |
| Female | 41 (50.0) | 17 (54.8) | 18 (72.0) | 0.153 |
| Disease duration, yr (n=133) | 2.23 [0.98, 3.97] | 2.95 [0.68, 5.29] | 1.19 [0.55, 3.40] | 0.362 |
| Interval between diagnosis and initiation of treatment (yr) | 0.12 [0.00, 0.54] | 0.35 [0.05, 1.23] | 0.13 [0.07, 0.41] | 0.108 |
| Clinical CNO lesion count* (n=61) | 1.00 [0.00, 2.75] | 1.50 [0.00, 3.50] | 1.00 [0.00, 2.00] | 0.861 |
| Total MRI lesion count* (n=43) | 5.00 [2.75, 8.25] | 4.50 [2.00, 5.00] | 4.00 [3.00, 9.00] | 0.909 |
| Active spinal lesion* | 6/25 (24.0) | 4/12 (33.3) | 8/17 (47.1) | 0.299 |
| C-Reactive Protein (mg/dL) (median [IQR]) (n=91) | 0.80 [0.80, 0.80] | 0.80 [0.80, 1.94] | 0.80 [0.80, 13.94] | 0.001 |
| ESR (median [IQR]) (n=100) | 9.00 [5.00, 16.00] | 11.00 [7.00, 29.50] | 15.00 [8.00, 36.00] | 0.141 |
| Presence of psoriasis | 3 (3.7) | 2 (6.5) | 1 (4.0) | 0.842 |
| Presence of IBD | 4 (4.9) | 4 (12.9) | 1 (4.0) | 0.304 |

Two patients with unclear dates of initiation of medication or diagnosis of CNO were excluded. \*Timing was within 90 days prior to initiation of medications, if there are multiple scores, the closest one to the initiation of medications was used. DMARD: disease modifying anti-rheumatic drug, IBD: inflammatory bowel disease, IQR: interquartile range, MRI: magnetic resonance imaging

### Development of the CNO CDAS

On February 12, 2021, the CARRA CNO workgroup annual meeting was held virtually with 36 participants (comprising 6 family representatives, 24 pediatric rheumatologists, 2 adult rheumatologists, and 4 researchers or research associates). Results from a literature review of existing disease assessment tools including PedCNO (24), CROMRIS (23), RAI-CROMRIS (22), and radiologic index for NBO (RINBO) (18) were presented. The group was divided into six breakout rooms to focus on specific measurements of CNO disease activity from three domains: physical exam findings, labot ratory findings, and patient reported outcomes or physician assessment. The following nine measurements were nominated by the group for consideration: total number of bone lesions with swelling and/or focal tenderness and/or warmth (clinical CNO lesion count), active joint count, ESR, CRP, hemoglobin, alkaline phosphatase, patient/parent global assessment, patient pain, and patient reported outcomes measurement information system (PROMIS) upper and lower extremity function. A ranking poll was carried out, and the three variables ranked the highest were patient pain, patient global assessment, and clinical CNO lesion count. Therefore, the CNO CDAS is calculated as a sum score encompassing these three variables.

### Validation of the CNO CDAS

From November 2021, external validation surveys to patients/parents and physicians were administered within CHOIR. We included children with a clinical diagnosis of CNO made by treating physicians who completed the survey and had available core variable values at the baseline and/or follow up visits. Standard of care including NSAIDs and/or second-line treatments were carried out. Degrees of difficulty of using limb/trunk/jaw, fatigue, depression, worry, self-perceived disease activity, improvement/worsening in disease activity, and perception of treatment effectiveness were reported. The construct validity of the CDAS was determined for three aspects and showed a significant correlation with patient/parent reported difficulty with using an affected body part, fatigue, sadness, worry, and physician global assessment (**Table 3**). There was a strong correlation between the CDAS and reported difficulty with using a limb, the back, or jaw (rho=0.65, p<0.001), with a weak correlation between the CDAS and reported fatigue, sadness, and worry (rho=0.24-0.39, p<0.001). Furthermore, the CDAS was significantly correlated with patient reported inactive disease, mildly active disease, moderately active disease, and severely active disease (rho=0.75, p<0.001) (**Figure 1**). It was noted that 25% of subjects had CDAS value of two or greater during inactive disease. The change of CDAS was significant in patients reporting disease worsening or improvement (p<0.001) while not significant in those reporting unchanged disease (p=0.526) (**Table 4**). The median change in the CDAS in patients reporting a significant improvement after treatment initiation was -2.5 (−5.8-(−0.3)). The CDAS increase in patients reporting significant worsening was 11 (4-12) (**supplement 3**). The CDAS significantly decreased after the initiation of second-line treatments from 12 (8-15.5) to 5 (3-12) (p=0.002) (**Table 5**).

**Table 3.** CDAS convergent and divergent validation using patient reported outcome

| *Variables* | *Estimates* | *CI* | *Correlation rho* | *P-value* |
| --- | --- | --- | --- | --- |
| Physician assessment (0-10) (n=809) | 0.3 | 0.27 – 0.33 | 0.76 | <0.001 |
| Difficulty to use limb/back/jaw (n=209) | 0.29 | 0.24 – 0.35 | 0.66 | <0.001 |
| Fatigue (n=208) | 0.15 | 0.10 – 0.20 | 0.37 | <0.001 |
| Sadness (n=207) | 0.17 | 0.09 – 0.24 | 0.35 | <0.001 |
| Worry (n=207) | 0.13 | 0.07 – 0.19 | 0.24 | 0.002 |

CI: confidence interval. Data for Physician assessment were between 8/2018 and 9/2022 whereas data for other parameters were only available from 11/2021.

**Table 4.** Correlation of CDAS changes with patient reported disease status change

| **Grouping** | **n** | **Median [IQR]** | **P-value\*** |
| --- | --- | --- | --- |
| Delta CDAS in the reported worsening group | 16 | 9.00[5.25,11.25] | <0.001 |
| Delta CDAS in the reported improved group | 64 | - 3.50[-6.00, -0.75] | <0.001 |
| Delta CDAS in the reported unchanged group | 8 | - 1.00[-3.00, 0.50] | 0.526 |

\*Wilcoxon signed rank test with continuity correction comparing to zero

**Table 5.** CDAS changes after second-line treatments

| **Variables** | **Before** | **After** | **p-value** |
| --- | --- | --- | --- |
| Patient pain VAS (n=101) | 4.00 [1.00, 7.50] | 2.00 [0.00, 7.00] | 0.155 |
| Patient/parent global assessment VAS (n=53) | 6.00 [3.00, 8.00] | 2.00 [1.00, 5.00] | <0.001 |
| Total number of clinically active lesions (n=126) | 1.00 [0.00, 2.00] | 0.00 [0.00, 1.00] | <0.001 |
| CNO CDAS (n=41) | 12.00 [8.00, 15.50] | 5.00 [3.00, 12.00] | 0.002 |

VAS: visual analogue scale, CDAS: clinical disease activity score

**Figure 1.**
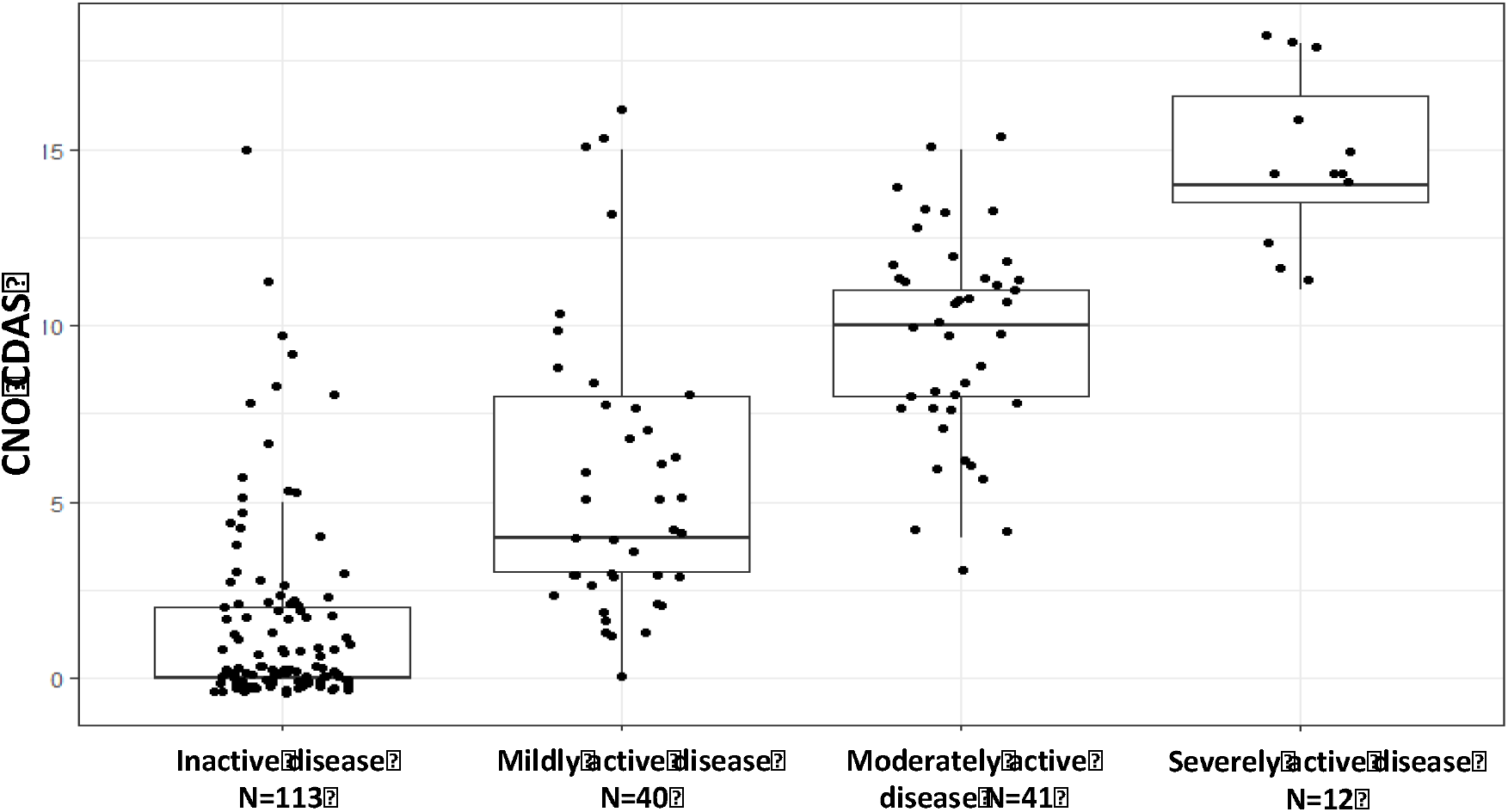
Plot of CDAS by the patient-reported disease severity. Spearman’s rank correlation showed rho =0.75, p<0.001.

### Response to CTP options

Of the 140 participants included in this study, 101 participants had complete data and 6-month follow-up. Due to the sequential exposure or concurrent administration of more than one CTP medication from baseline to follow-up, responses in clinical assessments were analyzed collectively. Treatment with a csDMARD, TNF inhibitor, and/or bisphosphonate resulted in significant improvement in clinical response (**Table 5**). Compared to baseline, patient pain (4 [1-8] versus 2 [0-7]) did not significantly differ, however, patient/parent global assessment of disease activity (6 [3-8] versus 2 [1-5]) and total number of clinically active lesions (1 [0-2] versus 0 [0-1]) all improved with second-line treatment (all p<0.05).

### Adverse events to CTP options

The total patient years for each CTP regimen were 351 for csDMARD, 294 for TNF inhibitors, and 89 for bisphosphonates. The most common adverse event was the development of psoriasis. There were 13 new cases of psoriasis, including two cases (among a total of 45 patient years) who developed psoriasis while taking leflunomide alone, seven cases (among a total of 114 patient years) while taking adalimumab, and four cases (among a total of 77 patient years) while taking infliximab. Two patients developed alopecia, one during treatment with leflunomide and the other with adalimumab. Two patients developed inflammatory arthritis, including one receiving a combination of pamidronate and methotrexate and one receiving methotrexate monotherapy. There was one patient each who developed a vertebral body compression fracture while on methotrexate, type 1 diabetes mellitus while on methotrexate, herpes zoster while on infliximab, and bacteremia requiring intravenous antibiotics while on infliximab and sub-therapeutic dose of methotrexate. Among reasons to discontinue a CTP medication, only two of 205 occasions were due to adverse events. Both patients discontinued infliximab due to severe psoriasis.

## Discussion

This pilot study demonstrated that the CARRA CNO CTPs are feasible and were used successfully across sites internationally within the CHOIR framework. The lack of significant differences in baseline patient characteristics between CTPs supports the diversity in prescribing patterns and allows the application of causal inference statistics to compare the effectiveness of non-NSAIDs treatments without randomization. Due to the rarity of the disease and the challenge of randomizing patients, and blinding patients and physicians from the treatments, large-scale randomized control trials have not been performed in children with CNO yet. Observational registries such as Eurofever (5), the German National Pediatric Rheumatology Registry (12), and now CHOIR can provide real world data to substantiate decision making for physicians and families. The use of biologics is limited in the two other registries but is enriched in CHOIR. A comparison of adalimumab and bisphosphonate has been proposed by a group of experts as a needed clinical trial (27). Recent retrospective cohort data suggest safety and efficacy of both, but higher relapse rates in patients treated with pamidronate (13). However, patients were not randomized and the cohort treated initially with TNFi exhibited higher inflammatory parameters and more bone lesions. Within the pamidronate group, failure to respond was associated with female gender, elevated systemic inflammatory parameters and increased numbers of bone lesions.

Results from this pilot study suggest that the CHOIR framework can be used to compare the effectiveness and safety among commonly prescribed medications. Notably, the distribution of characteristics of children with CNO receiving TNFi and those receiving bisphosphonates was comparable and balanced across groups, suggesting it is feasible to compare the effectiveness of these two medications within CHOIR. In a relatively large proportion of patients, there was a sequential use of csDMARD prior to use of TNFi or bisphosphonates, making it difficult to determine the effectiveness across all second-line treatments in CHOIR. However, retrospective studies (8,11) and surveys from families (28) suggest csDMARDs are less effective than TNFi and bisphosphonates for the treatment of CNO. Thus, a future main goal will be to focus on two second-line treatments used for CNO, TNFi and bisphosphonates, to provide more robust and accurate information as to their comparative safety and efficacy.

Prior to our study, there has not been any validated, standardized outcome assessment in CNO. The CNO CDAS has been validated through external questionnaires to patients/parents and physicians. Changes in CNO CDAS scores after second-line treatments were significant, while patient pain as an individual component did not reach significance. This may have been caused by the relatively high proportion of pain amplification in CNO (15,16), and highlights the comprehensive and robust nature of the CNO CDAS. The strong correlation coefficient between functional limitations with impaired use of arms/legs/back/jaw and physician global assessment with the CNO CDAS showed convergent validity, whereas the significant weak correlation between fatigue, depression, and worry and the CNO CDAS showed divergent validity. Furthermore, the excellent correlation of self-reported disease activity by patient/parent and the CNO CDAS established the capacity of the CNO CDAS as a disease severity assessment for future studies. While future study is needed to determine whether the CNO CDAS is a helpful measure that can be used not only in clinical research for CNO, but also in clinical practice, results promise potential for the CDAS score as a feasible and robust clinical-based disease activity assessment tool that can be calculated without requiring imaging data.

Safety monitoring has revealed psoriasis as the most common associated event within CHOIR, occurring in patients treated with leflunomide and monoclonal TNFi. Because psoriasis is highly associated with CNO, it is possible that a subset of patients developed psoriasis as part of natural history regardless of their exposure to these two types of medications. Nevertheless, it has been established that TNFi-associated psoriasis occurs in children with CNO (29–31), and the risk of paradoxical psoriasis after TNFi exposure is higher in children with CNO than in children with juvenile idiopathic arthritis or IBD (30). Concomitant use of methotrexate may decrease the risk of developing psoriasis (31,32). This is the first report of psoriasis occurring in a CNO patient treated with leflunomide, but further data is needed to determine if this is a new association. Other adverse events including the new onset of immune-mediated conditions and bacteremia were rare. There was no adverse event reported while patients received pamidronate suggesting its safety in children with CNO as previously reported (33). Overall, our data suggest that these medications have a favorable safety profile.

While providing promising data supporting the use of CHOIR and the new CDAS score to compare treatment outcomes in CNO, this study has several limitations. First, the observational nature of CHOIR does not allow randomization therefore the selection of treatments may be subject to the individual provider’s bias. However, the distribution of confounding factors is matched across different second-line treatments, which makes it possible to infer the relative effectiveness. Second, the follow up time is only two years for the inception cohort. Previous studies suggested that the median duration to a first disease flare ranges around 29 months in children treated with NSAIDs (11). However, we intend to continue data collection for 10 years or until patients transition to adult care. Third, missing data components of the CDAS from the visits prior to the validation of the CDAS did not allow for the comparison of treatment effectiveness using the CDAS. Future studies in CHOIR will include all of the components of the CDAS and lead to a clinical-score based assessment of disease activity and treatment response. Lastly, the MRI component of CHOIR has been collected based on site report of the total number of MRI lesions. A centralized imaging scoring will be needed to ensure that the readings of MRIs are reproducible.

Overall, this pilot study provided knowledge and experience that can be applied towards refining the CHOIR framework. Future efforts include building a larger cohort with more long-term data and clinical assessments to optimize analysis of the relative effectiveness of each CTP treatment option using the CDAS. We ultimately hope that results from larger-scale comparative effectiveness studies using this registry-based cohort can inform more robust evidence-based treatment guidelines and improve outcomes for this rare rheumatic disease.

## Conclusions

The CNO CDAS was developed and validated for disease monitoring and assessment of treatment effectiveness for children with CNO. CHOIR provides a feasible and comprehensive framework for comparative effectiveness research in the future.

## Data Availability

All data produced in the present study are available upon reasonable request to the authors

## Acknowledgements

The authors thank the CHOIR participants, research assistants and volunteers at all sites including Teresa Dickson, Corinne Lawler, Sumaya Aden, Thuan Bui, Kyra Shelton, Esha Mahal, Annie Xu, Kellen James, Shayla Nguyen, Zheng Xu, Ava Klein, Chessie Snider, Mabel Ho, Trang Pham, Anna Saack, Paige Trunnell, Emily Deng, Ana Park, Cailey Karshmer, Emma Leisinger, Mary Ellen Riordan, and Justine Griswold. We appreciate the help from Ingrid Goh, Mariana Correia Marques, and Min-Lee Chang for the testing of the registry database and the training material. In addition to the authors, the following CARRA CNO workgroup members participated in the February 2021 meeting: Ingrid Goh, Brian Nolan, Tzielan Lee, Annette Jansson, Aleksander Lenert, Lina Jaberi, David Cabral, Lauren Potts, Arielle Hay, Karine Toupin-April, Akaluck Thatayatikom, Ingram Chang, Piya Lahiry, Anja Schnabel, Mikhail Kostik, Nathan Rogers, Achille Marino, Dita Cebecauerova, Phillip Mease, Lindsey Bergstrom, Suzanne Li, Deborah McCurdy, Alex Theos, Matthew Hollander, Samira Nazzar, Farzana Nuruzzaman, Beverley Shea, and Chris Obrien.

---

**Supplement 1.**
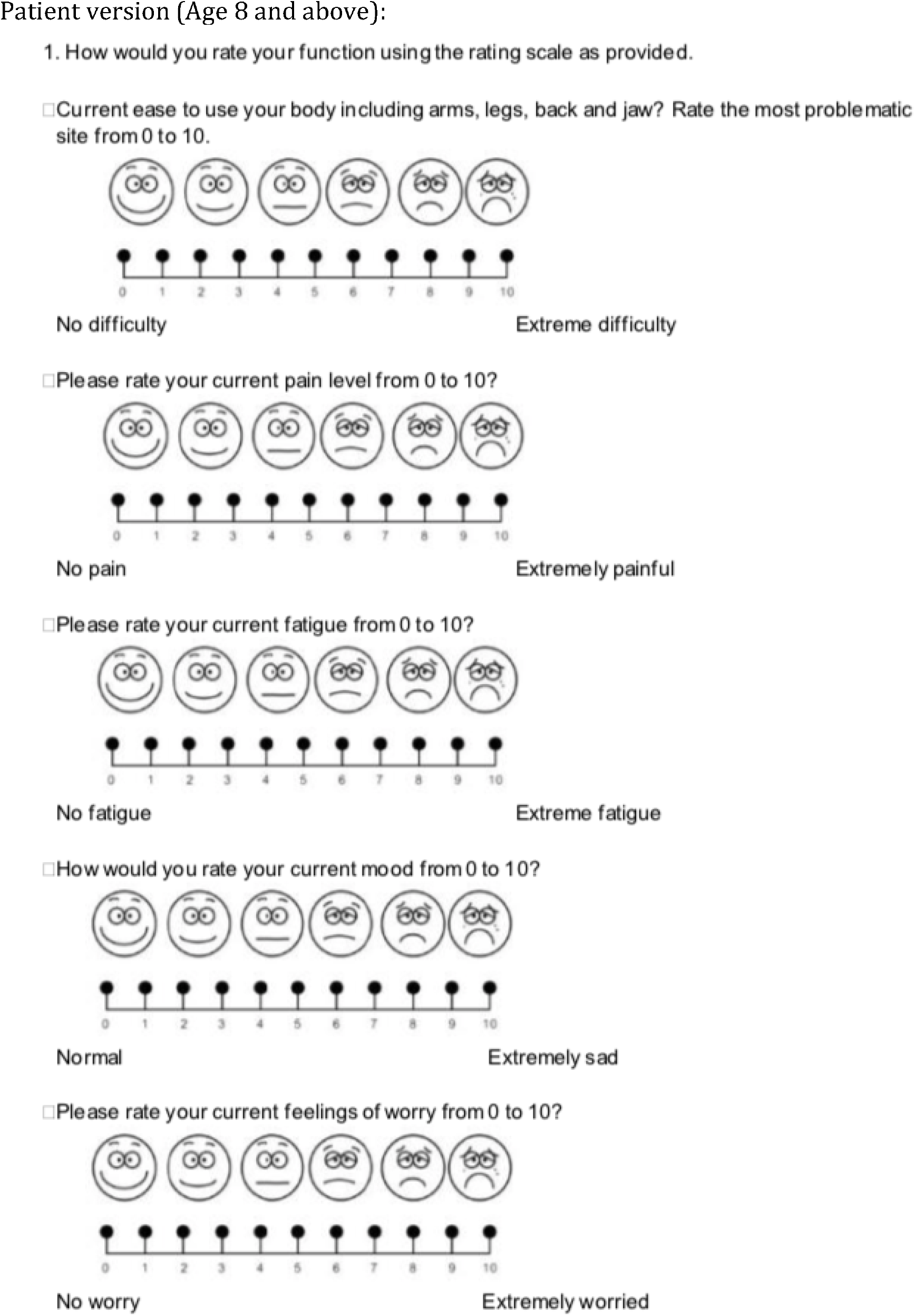
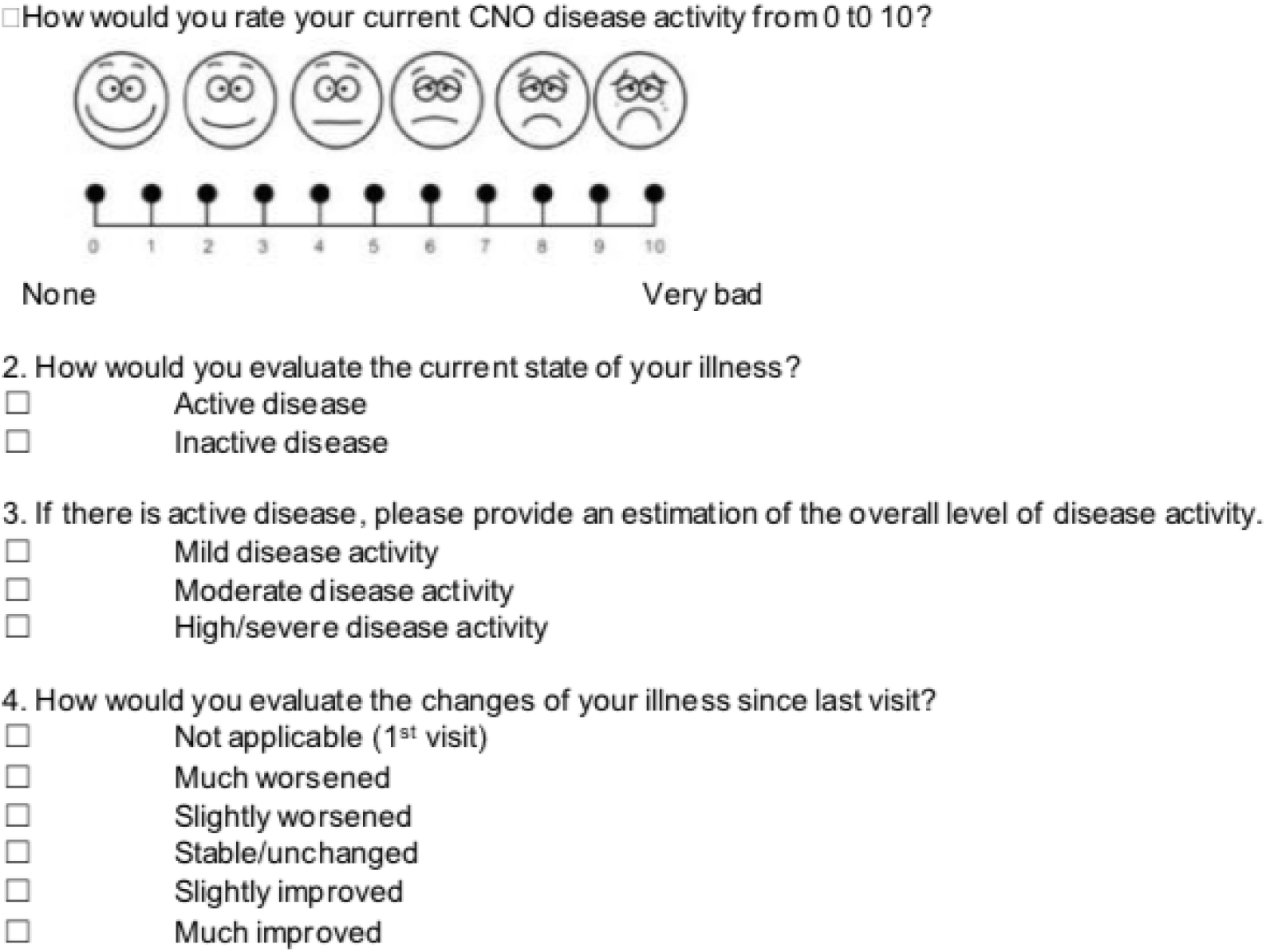
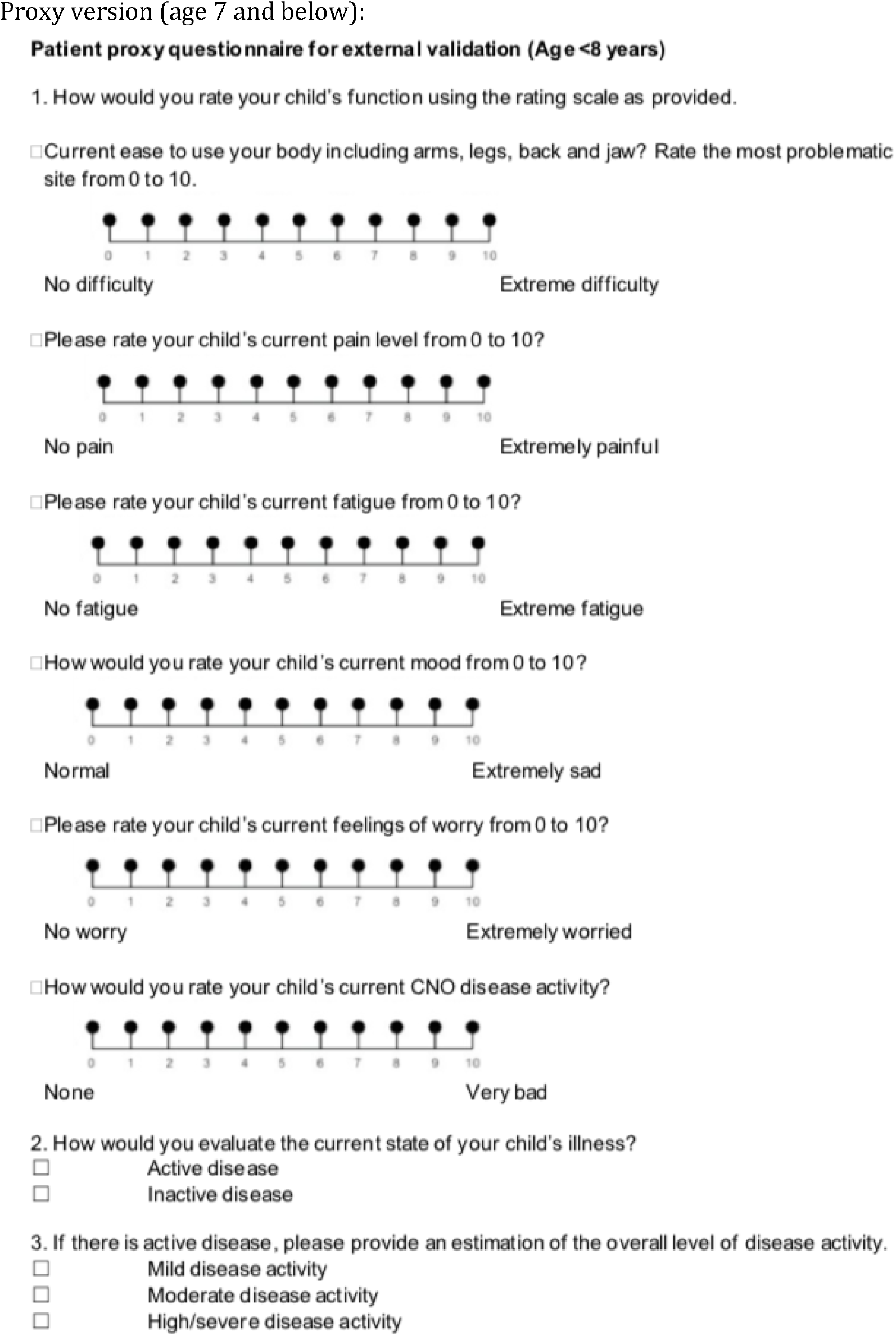
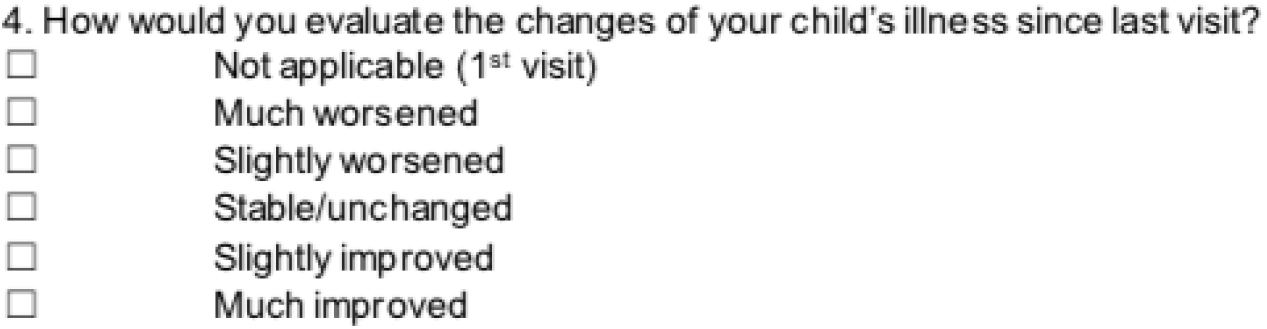
External validation surveys

**Supplement 2.**
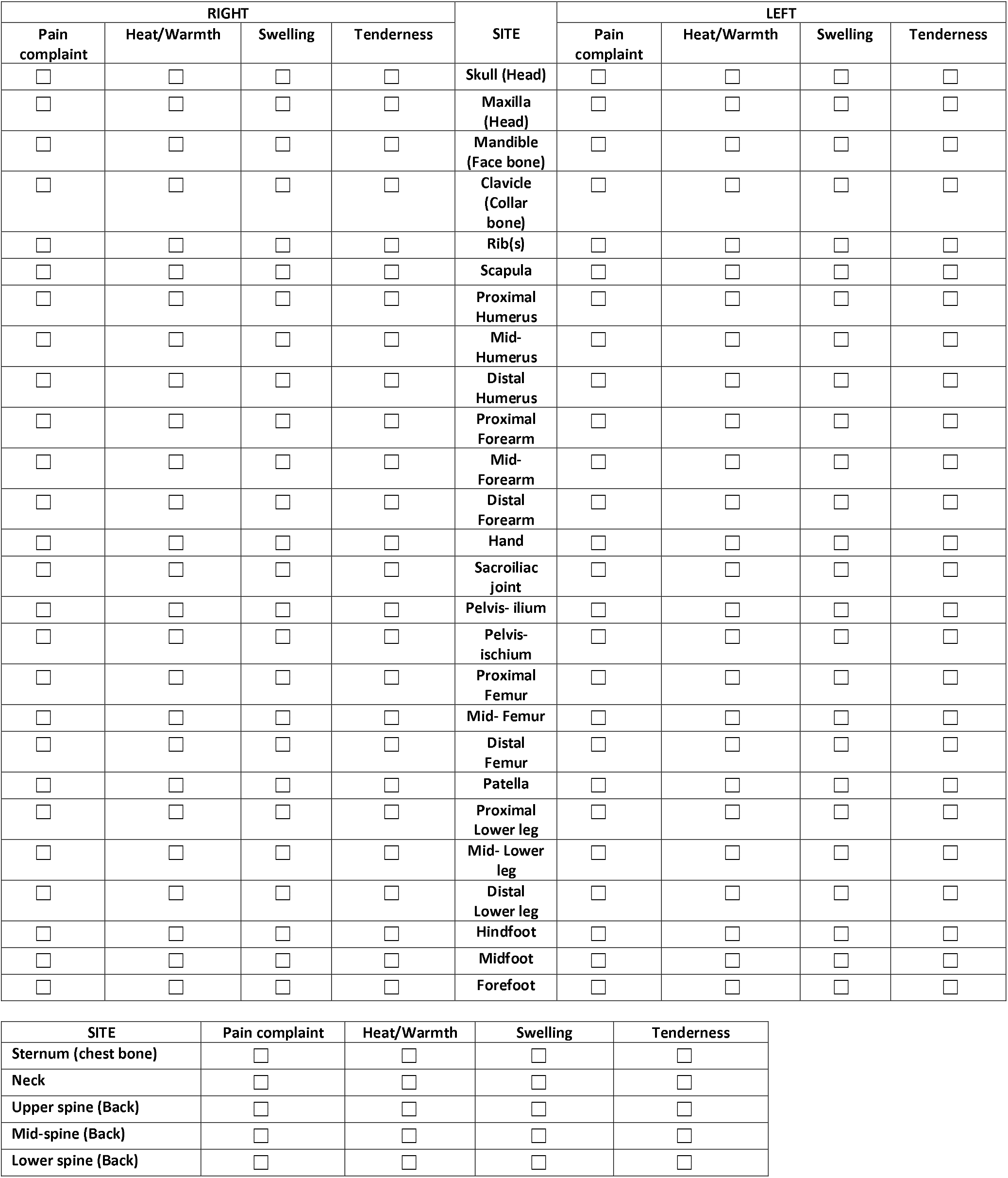
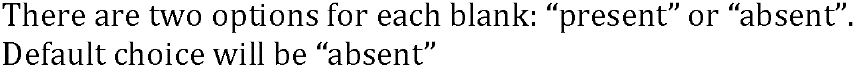
Physical exam recording sheet for children with CNO Please mark with ‘X’ physical exam findings:

**Supplement 3.**
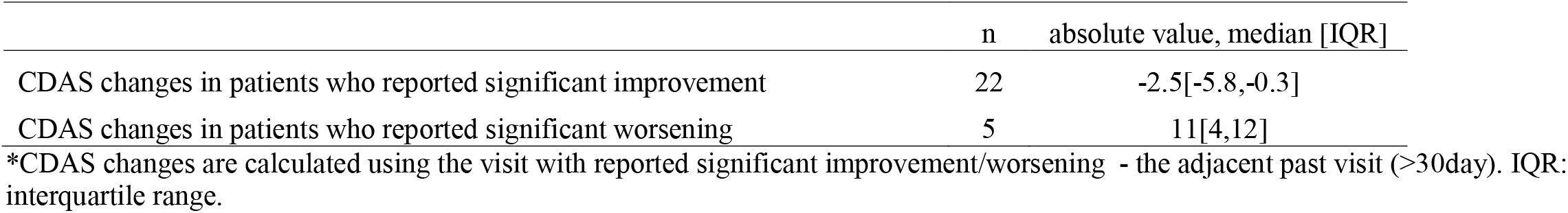
The changes of CDAS from groups who reported significantly improved or significantly worsened*

|  | n | absolute value, median [IQR] |
| --- | --- | --- |
| CDAS changes in patients who reported significant improvement | 22 | -2.5[-5.8,-0.3] |
| CDAS changes in patients who reported significant worsening | 5 | 11[4,12] |

\*CDAS changes are calculated using the visit with reported significant improvement/worsening - the adjacent past visit (>30day). IQR: interquartile range.

